# Temporal changes in pneumococcal carriage among childcare-attending children during the COVID-19 pandemic in the Greater New Haven Area, USA

**DOI:** 10.1101/2025.05.15.25327698

**Authors:** Chikondi Peno, Deus Thindwa, Maikel S. Hislop, Devyn Yolda-Carr, Damilola Egbewole, Giri Viswanathan, Tzu-Yi Lin, Sidiya Mbodj, Erica Rayack, Hibah Mahwish Askari, Yasaman Kazemi, Sarah Lapidus, Erica S. Spatz, Elissa Zirinsky, Carlos R. Oliveira, Amy K. Bei, Daniel M. Weinberger, Anne L. Wyllie

## Abstract

**Introduction:** Non-pharmaceutical interventions (NPIs), which aimed to reduce transmission of SARS-CoV-2, also reduced the circulation of other respiratory pathogens. However, it remains unclear to what extent NPIs disrupted pneumococcal colonization in childcare settings. We investigated pneumococcal colonization dynamics in childcare centers in the Greater New Haven Area (USA) with varying levels of COVID-19-related NPI recommendations.

**Methods:** Weekly, parent-collected saliva samples were obtained from children attending 8 childcare centers during Spring 2021 (February 2021-June 2021) and Winter/Spring 2021/22 (November 2021-June 2022). Samples were culture-enriched and tested using quantitative polymerase chain (qPCR) for pneumococcus targeting *piaB, lytA*, and pneumococcal serotypes. Trajectories of pneumococcal colonization were modeled using Markov models adjusted for age, sex, household size, ethnicity and study period. Bayes’ rule was used to adjust for false positivity of detecting non-pneumococcal signals in pneumococcal serotype-specific qPCR assays.

**Results:** A total of 1,100 saliva samples were collected from 100 children aged 0.3-5.9 years (median = 3.3 years, IQR = 1.8-4.6 years). Carriage dynamics based on detection of *piaB* and *lytA* were largely similar. Aggregated *piaB*-based carriage prevalence was higher during Winter/Spring 2021/22 (52.8%, 95% Confidence Interval: 49.4-56.3) than Spring 2021 (30.7%, 25.4-36.5). This increase in prevalence during Winter/Spring 2021/22 was driven by a higher proportion of high-density carriers, coinciding with less stringent NPI measures.

Overall carriage acquisition rates were also higher during Winter/Spring 2021/22 than Spring 2021 (Hazard Ratio [HR]: 2.88, 1.09-7.60). The most commonly prevalent serotypes were 15B/C, 11A/D/E and 33F/A/37.

**Conclusions:** We observed distinct patterns of pneumococcal carriage between Spring 2021 and Winter/Spring 2021/22 of the COVID-19 pandemic, among children attending New Haven childcare centers. The increase in carriage prevalence coincided with the relaxation of COVID-19 NPI measures during Winter/Spring 2021/22 and an increase in local cases of certain respiratory viruses.

## INTRODUCTION

*Streptococcus pneumoniae* (pneumococcus) is a common colonizer of the upper respiratory tract but is also a significant cause of both acute respiratory tract infections and invasive disease worldwide. While colonization is often asymptomatic, it is a prerequisite for pneumococcal disease and facilitates transmission between individuals in the population [1, 2]. Several factors, including young age, crowding, and viral co-infections, are well-established risk factors for pneumococcal carriage and subsequent disease [1].

Childcare centers play a key role in the transmission of pneumococcus [3]. Children in these settings are at a higher risk of acquiring pneumococcal carriage and subsequently developing pneumococcal disease [4, 5]. The crowded nature of childcare centers fosters person-to-person transmission of pneumococcus, and high environmental levels of viable pneumococci increase the risk of fomite transmission [6]. Subsequently, childcare attendees may transmit pneumococcus to more vulnerable household members, including younger siblings [7, 8], and older adults [9, 10]. Despite the important role of childcare centers in these transmission dynamics, studies on pneumococcal carriage in these settings remain limited.

The COVID-19 pandemic provided an opportunity to investigate how non-pharmaceutical interventions (NPIs) aimed at limiting SARS-CoV-2 transmission also affected the transmission of other respiratory pathogens. Starting from the early stages of the pandemic in March 2020, global reports showed a substantial reduction in infections caused by respiratory pathogens such as respiratory syncytial virus (RSV) [11], influenza viruses [12, 13] and invasive pneumococcal disease (IPD) [14, 15]. These declines coincided with widespread adoption of NPIs, including social distancing measures, mask-wearing, and limited in-person interactions. However, the extent to which these NPIs disrupted pneumococcal colonization and serotype distribution in childcare centers remains unclear, with conflicting pieces of evidence [16–18].

Therefore, during the spring of 2021 and the winter/spring season of 2021/2022, we investigated the impact of the COVID-19 pandemic on the longitudinal dynamics of pneumococcal carriage and the serotype distribution in children attending childcare centers in the Greater New Haven Area, (CT, USA).

## METHODS

### Ethical approval

The study was approved by the Institutional Review Board of the Yale Human Research Protection Program (Protocol number: 200002839) [19]. Parents or guardians provided written informed consent for their children to participate in the study.

### Study design and data collection

We conducted a prospective cohort study in children attending childcare centers in the Greater New Haven Area, CT, USA during Spring 2021 (February-June 2021) and Winter/Spring 2021/22 (October 2021-June 2022) [19]. Each week, saliva samples were collected from each study participant at home by their parent/guardian, as previously described [19]. In addition to sample collection, data on potential risk factors were collected at baseline and included age, race, ethnicity, sex, childcare center, household size, and presence of household members in different age strata. Individual vaccination records were unavailable during the study, but pneumococcal conjugate vaccine (PCV) vaccination coverage of three primary dose series (at 2, 4, 6 months-old) and with a booster dose (at 12-15 months-old) was reported at 92.3% in Connecticut in 2021 [20].

### Sample processing for pneumococcal detection

Saliva samples were dropped off at the study participant’s childcare center, collected by the study team, transported to the lab at ambient temperature, and stored at-70°C for further processing. Samples were culture-enriched for pneumococcus by inoculating 100 μL of raw saliva on trypticase soy agar supplemented with 7% defibrinated sheep blood and 10 mg/L gentamicin (SB7-GENT plates, ThermoFisher Scientific) and incubated overnight at 37°C with 5% CO_2_. Bacterial growth was harvested into 2100 μL of Brain Heart Infusion (Oxoid) supplemented with 10% glycerol and stored at-80°C until further analysis [9]. Bacterial DNA was extracted from 200 μL of the culture-enriched sample using a modified MagMAX Viral/Pathogen Nucleic Acid Isolation kit (ThermoFisher Scientific) protocol on a KingFisher Apex (ThermoFisher Scientific) [9].

### Detection of pneumococcus and serotype determination

Real-time quantitative polymerase (qPCR) using primers and probes specific for pneumococcal genes, *piaB* [21, 22] and *lytA* [23] were used to detect the presence of pneumococcal DNA [9]. Samples yielding a Ct-value of <40 were considered positive for pneumococcus. DNA from samples that tested positive for *piaB* were pooled by five whereas DNA from samples that tested negative were pooled by ten and tested in parallel with positive samples to evaluate the specificity of serotype-specific qPCR. Pooled DNA was tested in seven multiplexed serotyping RT-qPCR assays covering a total of 22 serotypes or serogroups (1, 2, 3, 6A/B/C/D, 7F/A, 8, 9L/N, 9V/A, 10A, 11A/D/E, 14, 15B/C, 19A, 19F, 20, 22F/A, 23A, 23B, 28F/A, 16F, 33F/A/37,34). Samples from pools testing positive on any multiplexed assay were re-tested individually on the corresponding singleplex assays to determine specific sample and serotype positivity.

### Respiratory virus detection

Samples were tested on the day of receipt for SARS-CoV-2 using an extraction-free, dualplex qRT-PCR assay (SalivaDirect) targeting the N1 gene and human RNase P (RP) as a sample quality control [19, 24]. An extraction-free, multiplex RT-qPCR protocol was additionally used to detect influenza A, influenza B, RSV and human metapneumovirus (hMPV), and human RNase P (RP) as sample quality control [25]. In brief, saliva lysates were prepared following the “SalivaDirect” extraction-free protocol by adding 6.5 µL (20 mg/mL) of proteinase K (New England Biolabs) to 50 µL of each saliva sample and heat-treated at 95°C for 5 minutes [24]. A 5 µl volume of the saliva lysate was used as a template for the multiplex RT-qPCR. A pooled positive control containing 100 RNA copies of each target (Twist Bioscience,CA) and a negative qPCR control were included in each run.

Samples were classified as positive when a Ct-value of <40 was detected [25].

In addition, we compared pneumococcal carriage trends detected in this study with publicly available weekly reports of respiratory illness activity, by analyzing data from Yale’s laboratory-based respiratory illness surveillance system [26]. This system captures respiratory viral test results from patients tested at Yale New Haven Hospital, regional hospitals, and community clinics. Samples were analyzed by the surveillance system using either a 4-plex PCR platform which detects influenza A, influenza B, SARS-CoV-2, and RSV or a respiratory virus PCR panel, which tests for influenza A & B, RSV A & B, human metapneumovirus (HMPV), parainfluenza types 1–4, adenoviruses, coronaviruses (229E, OC43, NL63, HKU1), and rhinoviruses.

### Statistical analyses

Descriptive analyses were used to characterize the study population by calculating the proportion of participants in each study period (Spring 2021 and Winter/Spring 2021/22), stratified by each independent potential risk factor described above. Samples were aggregated to monthly timescale to calculate observed pneumococcal prevalence stratified by qPCR target, study season and pneumococcal carriage density (high, *piaB* Ct-value <30 vs low, *piaB* Ct-value ≥ 30). Carriage prevalence was calculated as the monthly number of pneumococcal positive samples divided by all samples during that month.

We adapted previous models that have extensively been used for pneumococcal carriage dynamics of overall carriage (all serotypes combined) and individual serotypes, while adjusting for potential risk factors in a multivariate framework [27–30]. In brief, the Markov models for overall carriage were structured as having uncolonized (5) and colonized (1_l_, 1_2_) states, with associated rates of acquisition (A_l_, A_2_) and clearance (r_l_, r_2_) where subscripts 1 and 2 represent any two different serotypes. Individuals switching serotypes between the current and next time point without a clearance episode in between were allowed to transition at the rate (*ϕ*_1_λ_2_ or *ϕ*_2_λ_l_) where *ϕ*_l_(*ϕ*_2_) is the susceptibility of 1_l_(1_2_) carriers to switch to 1_2_(1_l_) relative to switching to uncolonized state. All parameters were constrained, such that λ_l_ = λ_2_, r_l_ = r_2_, and *ϕ*_l_ = *ϕ*_2_, allowing estimation of overall carriage acquisition and clearance without biasing duration of carriage from prolonged carriage of multiple subsequent serotypes (Fig 1A). Models for individual serotypes reflected independent serotype acquisition (λ_l_) and clearance (r_l_) rates between uncolonized and colonized states (Fig 1B).

**Figure 1.**
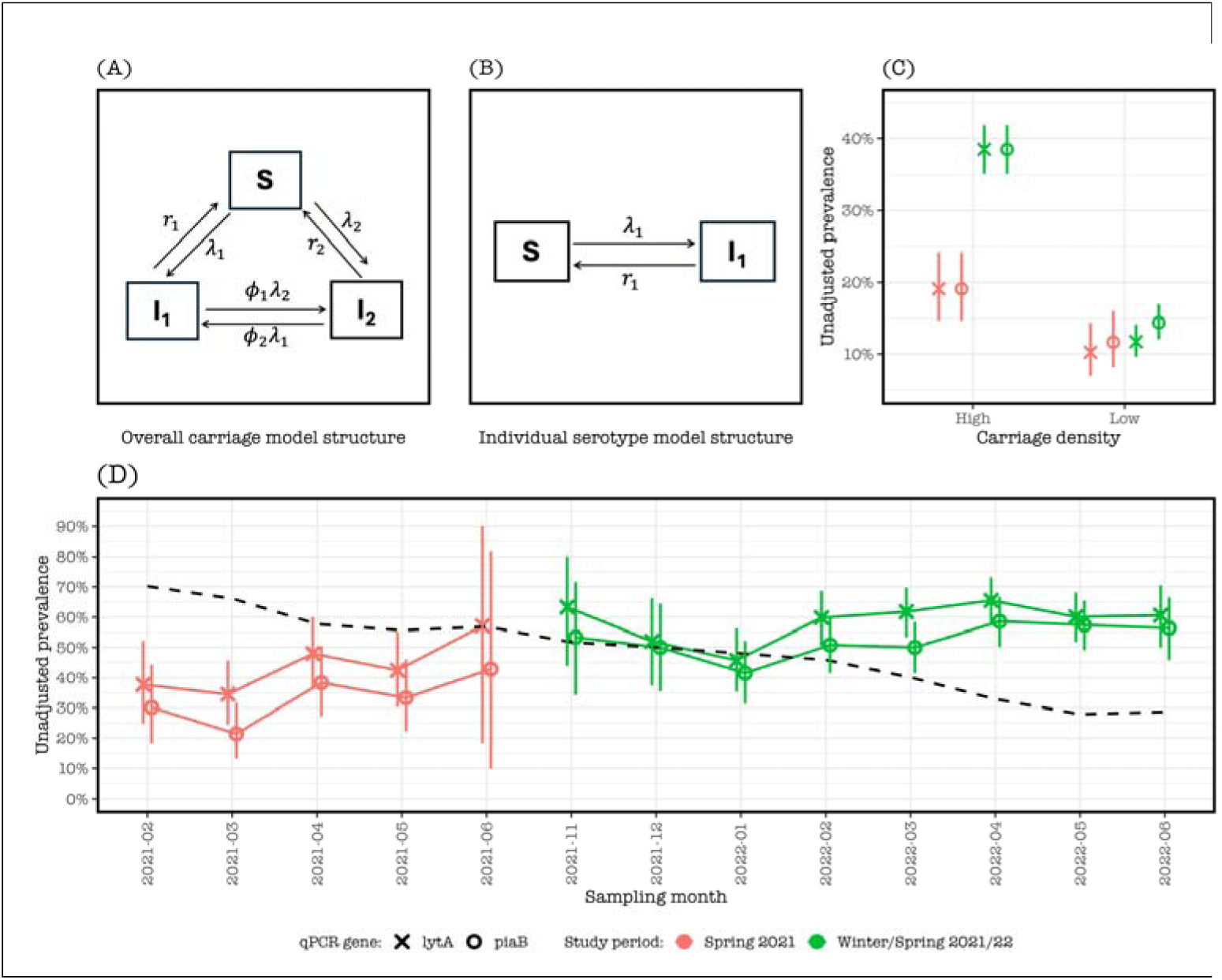
Pneumococcal colonization dynamics among children attending childcare centers in the Greater New Haven Area during February 2021-June 2021 (‘Spring 2021’) and November 2021-June 2022 (‘Winter/Spring 2021/22’). (A) Overall pneumococcal carriage model (all serotype combined) with states for uncolonized (S), current colonized (I_l_) and next colonized (I_2_), and acquisition rates (λ_l_, λ_2_) and clearance rates (r_l_, r_2_) of current and next carriage, and switch rates (*ϕ*_l_λ_2_, *ϕ*_2_λ_l_) rate between colonized states. (B) Individual serotype model with states (5, 1_1_) and serotype acquisition rate (λ_l_) and clearance rate (r_l_). (C) Higher observed pneumococcal carriage prevalence during Winter/Spring 2021/22 than Spring 2021 among children with high carriage density irrespective of pneumococcal detection method. High and low pneumococcal carriage densities were defined as a Ct-value <30 and ≥30, respectively, with all samples without detectable pneumococcal genes excluded. (D) Time series of monthly aggregates of observed pneumococcal carriage detection showing consistently higher *lytA-* than *piaB-based* carriage prevalence across study periods where the dashed black line is the US contact stringency index from the Oxford COVID-19 Government Response Tracker.

Models for both overall carriage and individual serotype were fitted to carriage datasets during Spring 2021, Winter/Spring 2021/22, and combined study periods. The time spent in the colonized state was assumed to be exponentially distributed such that carriage duration was an inverse of the carriage clearance rate. Acquisition and clearance rates were estimated by maximizing the log-likelihood. Individuals already carrying pneumococci at baseline were assumed to have acquired those pneumococci at a rate similar to steady state rates over the study period. Potential risk factors were only included in the model of the overall carriage outcome due to sample size limitation. The effect of risk factors on transition intensities was modelled using proportional intensities through hazard rates. Models were implemented using the ‘msm’ package in R software and optimized using bound optimization by quadratic approximation algorithm [31].

### Correcting for detection of pneumococcal serotype-specific qPCR assay false positivity

Given that serotyping was done by qPCR and that there was substantial detection of serotypes in samples negative for *piaB* and *lytA*, we used Bayes’ rule to estimate the probability that a positive detection by qPCR was a true pneumococcal serotype detection [32]. We assumed that the pre-test probability of detecting a pneumococcal serotype was a function of the reported number of IPD cases in the US in 2018-2021 and serotype-specific invasiveness e.g.,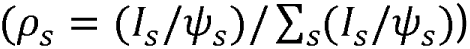 where ρ is the inferred serotype proportion in carriage, 1 is the reported number of IPD cases and *ψ* is the invasiveness of serotype s. The observed prevalence of each serotype in *piaB-*positive samples gave the false-positive rate, and we assumed that the test sensitivity was 100%. Thus, the probability that a sample testing positive for a specific serotype was truly positive for a pneumococcus expressing that serotype is given by 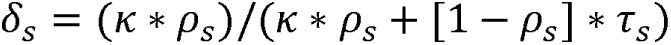 where δ is the probability that the serotype s sample is truly positive for pneumococcus, κ is the sensitivity of the *piaB* test set to 100%, p is the inferred serotype prevalence from the US population, and r is the serotype prevalence observed in the study and assumed to be false positive. The estimated probability o_S_ was then multiplied by the estimated acquisition rates of individual serotypes to obtain adjusted acquisition rates. Serotypes 7F/A, 9V/A, and 28F/A were not among serotypes that informed analysis of overall carriage due to their significant classification bias based on Bayes’ rule, signifying high false positivity probability. Additionally, only *piaB* carriage patterns were modelled given the similarity with *lytA* results.

## RESULTS

### Participant characteristics

A total of 1,110 saliva samples were collected from 100 children (median age = 3.3 years, interquartile range [IQR]: 1.8-4.6 years) attending eight childcare centers in the Greater New Haven Area (**Table 1**). Of all saliva samples collected, 283 samples (25.5%) were collected from 27 children during the Spring 2021 period and 827 (74.5%) were collected from 73 children during the Winter/Spring 2021/22 period. Nine children (9%) participated in both study periods. The number of children sampled in each childcare center varied between the two study periods, with three childcare centers added to the study during the Winter/Spring 2021/22 study period.

**Table 1:**
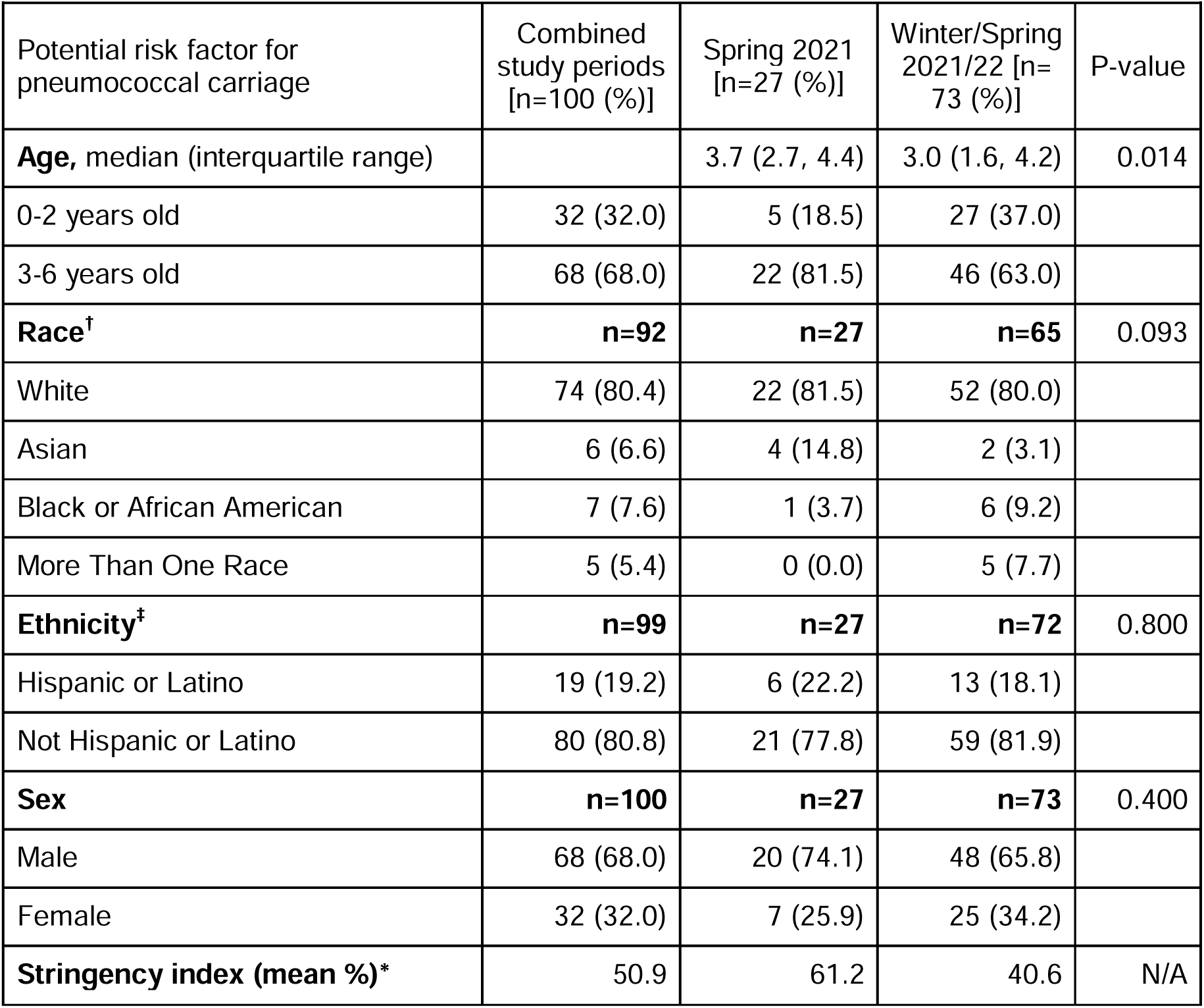
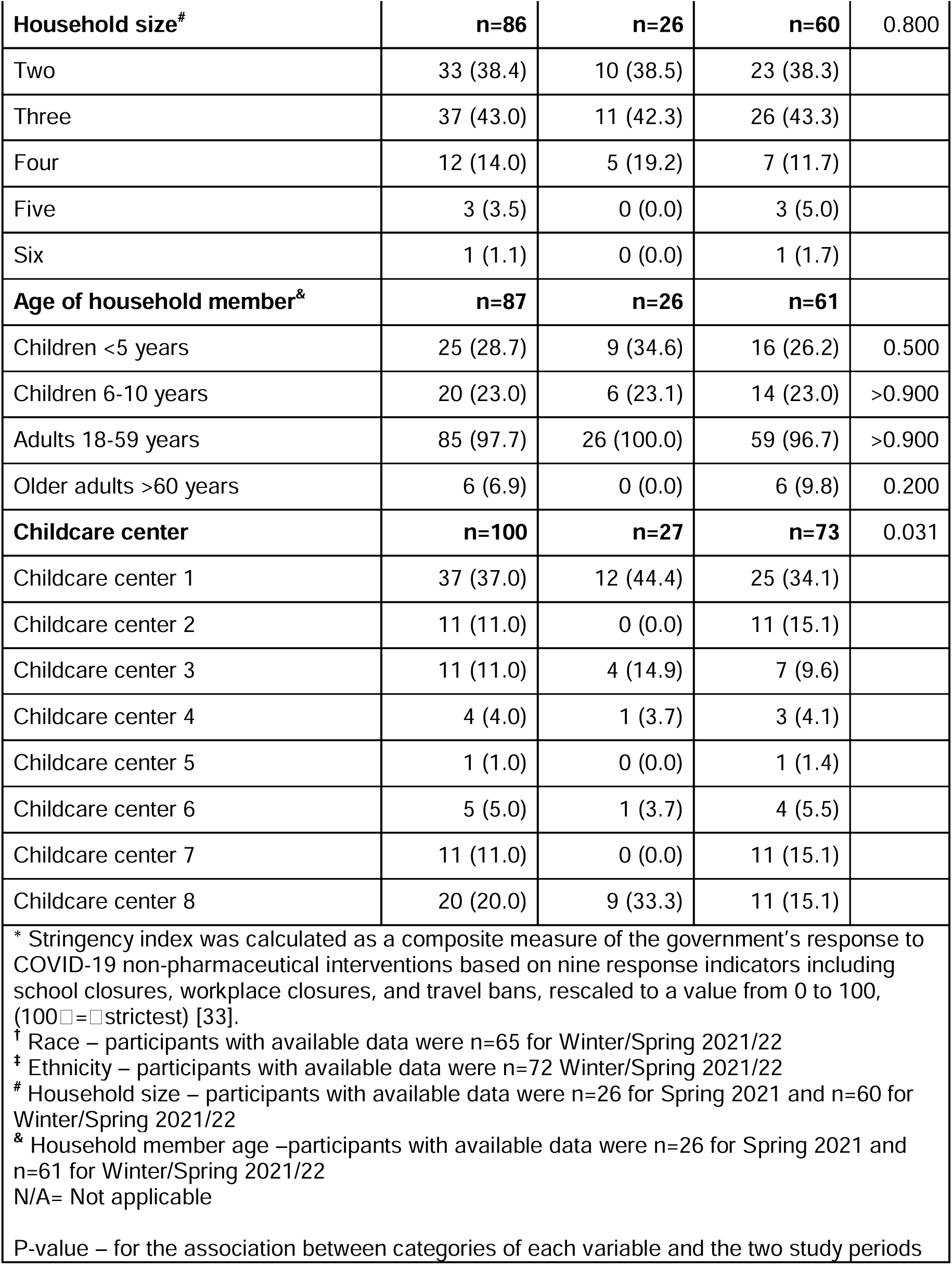
Baseline characteristics of the study participants overall and for each of the two study periods (Spring 2021, February-June; Winter/Spring 2021/22, October 2021-June 2022).

Children recruited in Spring 2021 were older than those recruited in Winter/Spring 2021/22 periods (*p*=0.014). Participants were predominately of White race in both study periods (81.5%, Spring 2021 vs. 80.0%, Winter/Spring 2021/22), and of non-Hispanic/Latino ethnicity (77.8%, Spring 2021 vs. 81.9%, Winter/Spring 2021/22). All other baseline characteristics did not significantly differ between the two study periods (**Table 1**).

### Observed pneumococcal carriage prevalence

Overall, 524/1100 (47.2%) saliva samples collected over the study tested positive for *piaB,* 607 (54.7%) samples tested positive for *lytA* and 498 (44.9%) samples tested positive for both *piaB* and *lytA*. The increase in carriage prevalence in Winter/Spring 2021/22, was mostly seen for higher carriage density (Ct <30) as compared to lower carriage density (Ct≥30) (Fig 1B). Irrespective of study period, monthly detection of *lytA* was higher than *piaB* (range: 34.5-65.5% vs. 21.4-58.6%, respectively), likely reflecting the presence of non-pneumococcal Streptococci. Aggregated pneumococcal carriage prevalence based on the detection of *piaB* was higher during Winter/Spring 2021/22 than Spring 2021 (52.8%, 49.4-56.3 vs. 30.7%, 25.4-36.5, respectively). Similarly, the frequency of detection of *lytA* was higher during Winter/Spring 2021/22 than Spring 2021 (59.4%, 55.9-62.7 vs. 41.0%, 35.2-47.0, respectively) (Fig 1C).

### Pneumococcal carriage acquisition and clearance rates

Daily pneumococcal acquisitions based on *piaB* were higher during Winter/Spring 2021/22 compared to Spring 2021 (Hazard Ratio [HR]: 2.88; 95%CI: 1.09-7.60) for carriage in combined study periods. Acquisition rates did not vary significantly between Spring 2021/22 and Winter 2021/22 seasons (HR:0.82, 95%CI: 0.43-1.57). However, in contrast to the Winter/Spring 2021/22 period, carriage acquisition during Spring 2021 was lower for male vs female (HR:0.21, 95%CI: 0.05-0.87) and for household with >2 members vs 2 members (HR:0.10, 95%CI: 0.04-0.28). The clearance rate was slower among 3-6 years old vs 0-2 years old children for carriage in combined study periods (HR:0.41, 95%CI: 0.19-0.89) and during Winter/Spring 2021/22 (HR:0.24, 95%CI: 0.07-0.78), corresponding to the average duration of carriage of 2.4 days (95%CI: 1.1-5.3) and 4.2 days (95%CI: 1.3-14.3), respectively (Table 2).

**Table 2:** Potential factors associated with daily pneumococcal carriage acquisition and clearance based on *piaB* quantitative polymerase chain reaction (qPCR) test.

| Pneumococcal dynamics | Hazard Ratio (95% Confidence Intervals) |  |  |
| --- | --- | --- | --- |
|  | Combined study periods<sup>†</sup> | Spring 2021<sup>†</sup> | Winter/Spring 2021/22<sup>†</sup> |
| **Pneumococcal acquisition** |  |  |  |
| **Age** |  |  |  |
| $\leq 2$ years old | Reference | Reference | Reference |
| 3-6 years old | 0.80 (0.31, 2.09) | 1.21 (0.12, 12.50) | 0.53 (0.20, 1.40) |
| **Sex** |  |  |  |
| Female | Reference | Reference | Reference |
| Male | 0.48 (0.20, 1.11) | 0.21 (0.05, 0.87)* | 1.10 (0.49, 2.46) |
| **Household size** |  |  |  |
| 2 members | Reference | Reference | Reference |
| >2 member | 0.48 (0.20, 1.14) | 0.10 (0.04, 0.28)* | 0.86 (0.54, 1.40) |
| **Ethnicity** |  |  |  |
| Hispanic | Reference | Reference | Reference |
| Non-Hispanic | 0.74 (0.34, 1.65) | 0.44 (0.11, 1.66) | 0.90 (0.51, 1.60) |
| **Seasonality** |  |  |  |
| Spring | Reference | - | Reference |
| Winter | 0.82 (0.43, 1.57) | - | 0.85 (0.54, 1.36) |
| **Study period** |  |  |  |
| Spring 2021 | Reference | - | - |
| Winter/Spring 2021/22 | 2.88 (1.09, 7.60)* | - | - |
| **Pneumococcal clearance** |  |  |  |
| **Age** |  |  |  |
| ≤ 2 years old | Reference | Reference | Reference |
| 3-6 years old | 0.41 (0.19, 0.89)* | 0.35 (0.11, 1.22) | 0.24 (0.07, 0.78)* |
| **Sex** |  |  |  |
| Female | Reference | Reference | Reference |
| Male | 0.60 (0.30, 1.17) | 0.30 (0.10, 0.92) | 1.69 (0.56, 5.09) |
| † Acquisition estimates are adjusted for age, sex, ethnicity, household size in Spring 2021 model, and additionally for seasonality in Winter/Spring 2021/22 model, and additionally for study period in 'combine study periods' model. Clearance estimates are adjusted for age and sex for all the three models. |  |  |  |
| * Statistical significance at 95% confidence intervals |  |  |  |

### Pneumococcal serotype dynamics

Of the 846 combined isolates in Spring 2021 and Winter/Spring 2021/22, eighteen serotypes were identified from 324 (38.2%) *piaB* positive samples. The adjusted prevalence of serotypes and serogroups was (in order) 15B/C (10.7%), 11A/D/E (7.8%), 33F/A/37 (6.5%), 19F (5.2%), 10A (4.0%), 23B (3.1%), 17 (2.5%), 16F (2.5%), 3 (2.2%), 23F (2.0%), 19A (1.6%), 23A (1.5%), 21 (1.0%), 22F/A (1.0%), 28F/A (0.7%), 7F/A (0.2%), 9V/A (0.1%), and 20 (0.1%). Serotypes 23A and 23B were only identified during Spring 2021, whereas 9V/A, 16F, 3, 21, 19F, 20, and 19A were only identified during Winter/Spring 2021/22 (Fig 2A). Based on adjusted carriage of combined study periods, annual serotype acquisitions per child were highest for serotype 33F/A/37 (1.9 episodes, 95%CI: 1.4-2.4), 19A (1.25, 0.4-3.6), 15B/C (1.15, 0.7-1.9) and 23B (1.1, 0.4-3.1). Acquisition rates of common serotypes between Spring 2021 and Winter/Spring 2021/22 did not differ significantly. The estimated average duration of carriage in combined study periods was longest for serotype 22F/A (101 days, 95%CI: 44-227) and 33F/A/37 (50 days, 95%CI; 28-91), and shortest for serotype 20 (3 days, 95%CI;1-8) and 23B (4 days, 95%CI; 2-11) (Fig 2B).

**Figure 2.**
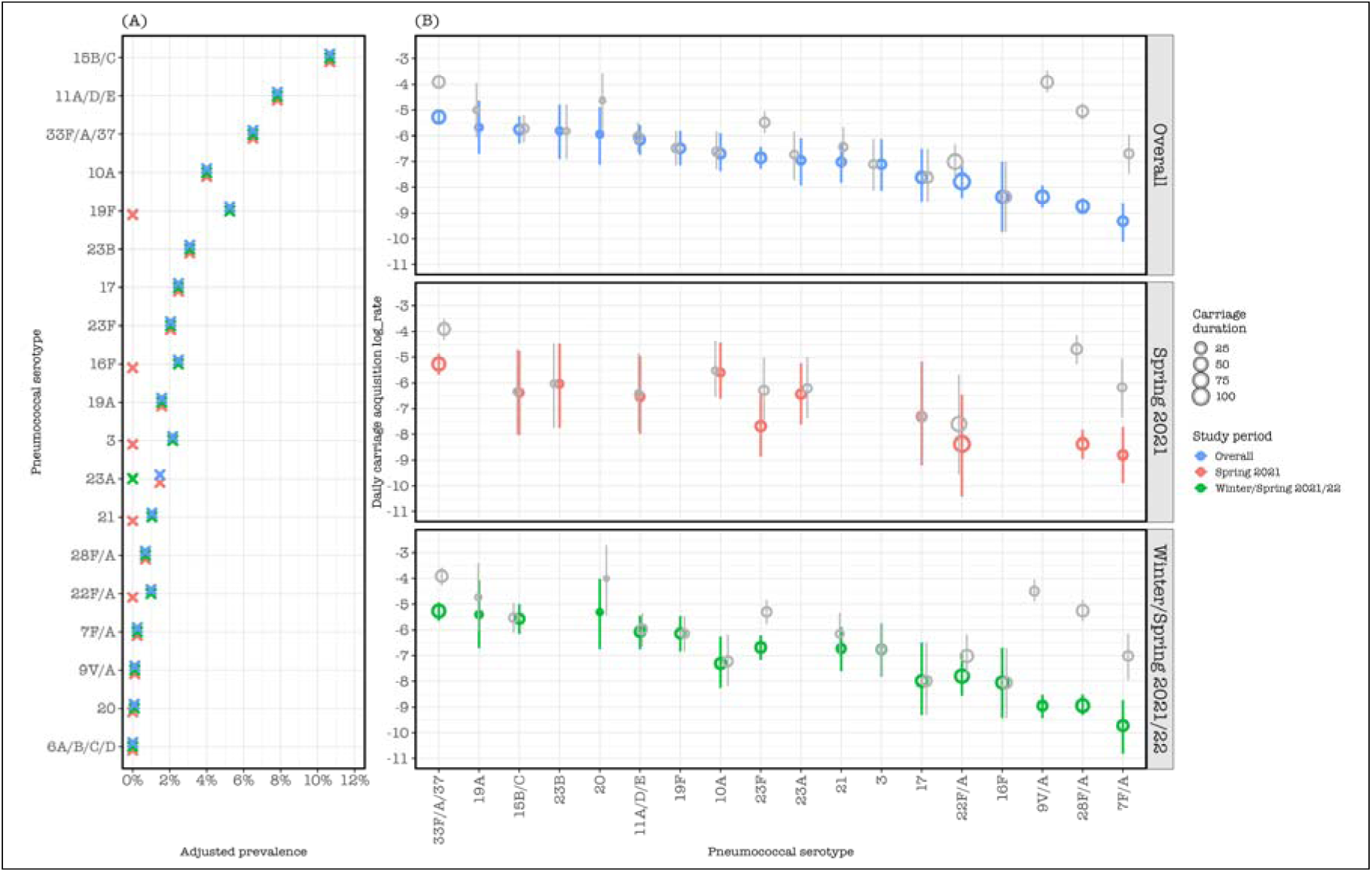
Serotype prevalence based on *piaB* positivity, with acquisition rates stratified by study period. (A) Serotype prevalence stratified by study period, with raw prevalence and prevalence based on correction for detecting non-pneumococcal Streptococci in the study (adjusted). (B) Study period stratified serotype acquisition rate based on unadjusted carriage in the study (gray circles) and based on adjusted carriage in Spring 2021 (red circles), in Winter/Spring 2021/22 (green circles) and in both Spring 2021 and Winter/Spring 2021/22 (blue circles). The vertical line across each circle represents 95% confidence interval of the serotype carriage acquisition point estimate. The size of the circle corresponds to the duration of serotype carriage. Both acquisition rate and carriage duration of each serotype are estimated using multistate Markov models. Only carriage acquisitions and duration of 18 serotypes for overall and Winter/Spring 2021/22 and 12 serotypes for Spring 2021 were estimated due to few acquisition events.

### Pneumococcal carriage prevalence relative to respiratory virus activity

Of the 1,110 saliva samples collected, 824 (74.2%) had adequate volume for viral testing. During Spring 2021, no sample tested positive for SARS-CoV-2, and 1 sample tested positive for influenza A virus. During Spring/Winter 2021/22, 1 sample tested positive for SARS-CoV-2, 2 samples tested positive for RSV and 1 sample tested positive for hMPV.

Since childcare center state policies required symptomatic children to isolate at home [34], respiratory virus activity in our study population may have been under detected. Comparison of our observations of pneumococcal carriage to the respiratory virus surveillance data reported by YNHH was performed to gain insight on respiratory viral activity in the area [26]. Pneumococcal carriage prevalence was relatively low during Spring 2021, consistent with YNHH virus testing data of all viruses. The increased prevalence and density in pneumococcal carriage that we observed during Winter/Spring 2021/22 reflected the dynamics of hMPV, parainfluenza viruses, rhinoviruses, and SARS-CoV-2 and coincided with relaxation of NPI measures. Cases of influenza A and B, RSV and other seasonal coronaviruses remained low during this period (Fig 1, Fig 3).

**Figure 3:**
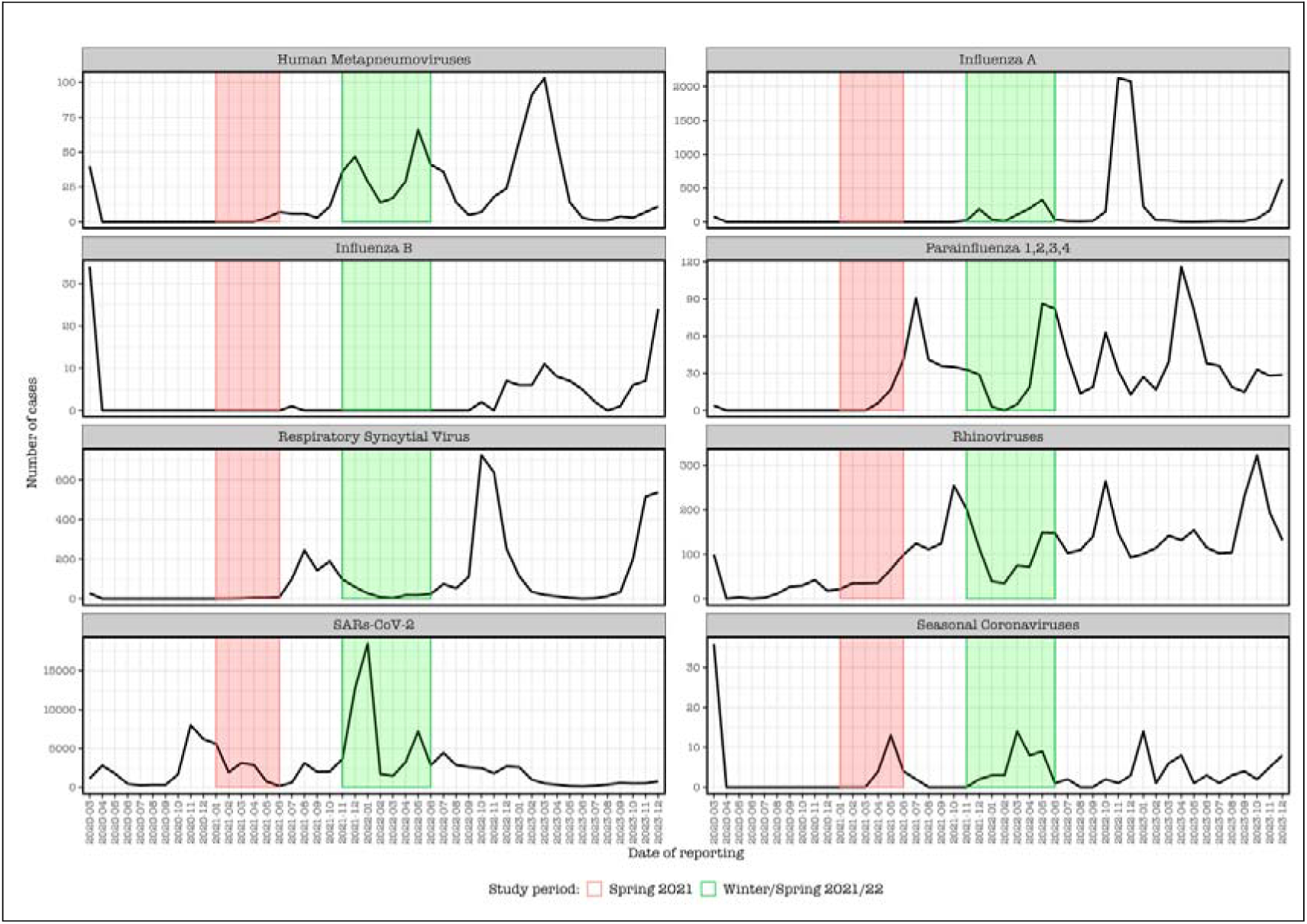
Weekly reports from Yale-New Haven Hospital (CT, USA) of certain respiratory virus cases from 2020-2023. The red and green shaded areas, respectively, correspond to the sampling periods (Spring 2021 and Winter/Spring 2021/22) of the current study, in which we investigated pneumococcal carriage dynamics from children in local daycare centers. The plots show substantial number of reported cases caused by SARS-CoV-2, influenza A and respiratory syncytial virus (RSV). It further shows fewer cases across all viruses during the spring of 2021 when social contact measures were more stringent, whereas during the Winter/spring of 2021/22, high number of cases of human metapneumovirus, parainfluenza viruses, rhinoviruses, and SARS-CoV-2 were reported, while cases of influenza A and B, RSV and other seasonal coronaviruses remained low.

## DISCUSSION

While children attending childcare centers are recognized as important reservoirs for respiratory pathogens, and stringent NPIs were in place during the COVID-19 pandemic to limit the spread of SARS-CoV-2, the extent to which pneumococcal carriage dynamics were disrupted in these settings remains unclear. Our longitudinal study provides insights into these dynamics, highlighting significant changes in pneumococcal carriage dynamics among attendees of childcare centers in the Greater New Haven Area (CT, USA) during two consecutive seasons of the COVID-19 pandemic. The higher rates of pneumococcal acquisition, prevalence and colonization density observed during Winter/Spring 2021/22 (November 2021-June 2022) compared to Spring 2021 (February 2021-June 2021), suggests a temporal influence of COVID-19 NPIs on pneumococcal carriage dynamics.

Specifically, we observed nearly 3-fold increase in overall pneumococcal carriage prevalence from Spring 2021 to Winter/Spring 2021/22, as well as higher density carriage (Ct<30) in Winter/Spring 2021/22. This shift in acquisitions rates might reflect the effect of relaxing NPIs of the COVID-19 pandemic [33]. Nonetheless, carriage prevalence levels remained high in both study periods. While we lack pre-pandemic data, studies in Belgium [17], Israel [16], Vietnam [35] and South Africa [18] have also reported sustained pneumococcal carriage among children over the course of the COVID-19 pandemic. In Vietnam [35] and South Africa [18], carriage densities were also reported to be lower during the pandemic than the pre-pandemic period[36]. The unexpected findings of lower carriage acquisition rates in households with more siblings and slower carriage clearance among older children should be cautiously interpreted as the limited number of children enrolled in this study likely skewed data on risk factors and subsequent inferences (Table 1). Possible explanations however could be due to school closures and less pneumococcal transmission among older siblings and, as suggested by our data, a more limited circulation of pneumococcal serotypes resulting in less inter-serotype competition in the carriage dynamics observed.

NPI measures aimed at COVID-19 containment are reported to have impacted circulation of other respiratory viruses including influenza and RSV, which in turn could have altered pneumococcal carriage pattens due to the known impact of respiratory viruses on pneumococcal carriage dynamics [37, 38]. During Spring 2021, our observation of lower density pneumococcal carriage prevalence was associated with stricter adherence to NPIs and low-level circulation of viruses. During Winter/Spring 2021/2021 however, NPIs were relaxed. Here, the increase in pneumococcal carriage prevalence coincided with increased circulation of human metapneumoviruses, parainfluenza’s, rhinoviruses and SARS-Cov-2 suggesting either potential interactions or a common risk factor (increased interaction in the population). Expectedly, less than 1% of carriage samples among healthy children were positive for the tested viruses, which are commonly isolated among symptomatic disease [39], and which may have precluded children from attending their childcare centers and thus providing samples during that period.

Pneumococcal carriage patterns and density for *piaB* and *lytA* were largely similar. However, our molecular serotyping assays may have detected some serotype-specific genes in samples negative for pneumococcus but rather containing non-pneumococcal Streptococci [40]. Our Bayes approach which showed that detection of serotypes 9V/A, 28F/A, 7F/A, 33F/A/37 and 23F were biased upward provides a useful method for dealing with analysis of false positivity in future studies. Although we observed more serotype diversity in the Winter/Spring 2021/22 period when NPIs were relaxed and both carriage prevalence and density increasing, our serotype acquisition analysis stratified by study period showed similar serotype-specific acquisitions rates for all tested serotypes in Spring 2021 and Winter/Spring 2021/22, possibly due to insufficient sample size.

A major strength of this study is its longitudinal design, which allowed us to quantify pneumococcal acquisition and clearance rates over time while accounting for individual carriage dynamics and seasonal variations, which are typically absent in cross-sectional studies. Parents being able to collect their children’s saliva was critical to the implementation of our study design, due to pandemic-related restrictions. Importantly, detection of pneumococcal carriage in saliva generally reflects rates detected by nasopharyngeal swabs [36]. Moreover, the combined use of *piaB* and *lytA* for the molecular detection of pneumococcus enhances specificity, particularly in saliva samples, where the presence of closely related *Streptococcus* species can confound single-gene assays [40, 41].

Several limitations should be noted. Most notably, we lacked pre-pandemic baseline data on pneumococcal prevalence in this population, which limits our ability to fully evaluate the impact of NPIs on carriage dynamics. In addition, since we were only able to enroll a subset of attendees from each childcare center, we were unable to definitively infer whether transmission events occurred within childcare centers or arose from community-based sources. Despite these limitations, our Bayes rule approach effectively discounted results from less reliable serotyping assays, and the 2-state colonization Markov approach of overall carriage reduced the carriage duration bias caused by prolonged carriage episode from multiple subsequent serotypes without clearance event in between.

In conclusion, our study reveals distinct patterns of pneumococcal carriage among childcare centers in the Greater New Haven Area during different periods of the COVID-19 pandemic. Despite high levels of pneumococcal carriage across both study periods, the relaxation of NPIs in Winter/Spring 2021/22 appears to have contributed to increased pneumococcal acquisition and prevalence. Observations from our study offer insight into the complex interplay between respiratory pathogens and the impact of NPIs, and the need for further research in more controlled/balanced/representative populations to fully elucidate the factors driving pneumococcal transmission in childcare settings.

## AVAILABILITY OF DATA AND MATERIALS

De-identified datasets used and/or analyzed during the current study are available from Github at https://github.com/weinbergerlab/spn.childcare.colonisation.usa

## AVAILABILITY OF CODE

The R code that was used for the study analysis can be accessed through Github at https://github.com/weinbergerlab/spn.childcare.colonisation.usa

## AUTHOR CONTRIBUTIONS

ALW conceived the study. DMW and ALW designed the study protocol. CP, AKB and ALW managed the study. CP, MH, DYC, DO and GV conducted laboratory experiments. CP, MH and DT analyzed the data. CP, DT, DMW and ALW drafted the manuscript. All authors amended, critically reviewed, and commented on the final manuscript.

## Data Availability

De-identified datasets used and/or analyzed during the current study are available from Github

https://github.com/weinbergerlab/spn.childcare.colonisation.usa

## ACKNOWLEDGEMENTS

We thank the study participants and their families for their time and dedication to the study. We thank all members of the Trackcare study team, Yale Pathology Labs and the Wyllie Lab at the Yale School of Public Health for their critical contributions to the research study

